# Brain functional network integrity sustains cognitive function despite atrophy in presymptomatic genetic frontotemporal dementia

**DOI:** 10.1101/19012203

**Authors:** Kamen A. Tsvetanov, Stefano Gazzina, P. Simon Jones, John van Swieten, Barbara Borroni, Raquel Sanchez-Valle, Fermin Moreno, Robert Laforce, Caroline Graff, Matthis Synofzik, Daniela Galimberti, Mario Masellis, Maria Carmela Tartaglia, Elizabeth Finger, Rik Vandenberghe, Alexandre de Mendonça, Fabrizio Tagliavini, Isabel Santana, Simon Ducharme, Chris Butler, Alexander Gerhard, Adrian Danek, Johannes Levin, Markus Otto, Giovanni Frisoni, Roberta Ghidoni, Sandro Sorbi, Jonathan D. Rohrer, James B. Rowe, on behalf of the Genetic FTD Initiative, GENFI

**Affiliations:** Department of Clinical Neurosciences, University of Cambridge, Cambridge, UK; Cambridge Centre for Ageing and Neuroscience (Cam-CAN), University of Cambridge and MRC Cognition and Brain Sciences Unit, Cambridge, UK; Department of Neurology, Erasmus Medical Center, Rotterdam, The Netherlands; Centre for Neurodegenerative Disorders, Neurology Unit, Department of Clinical and Experimental Sciences, University of Brescia, Brescia, Italy; Alzheimer’s disease and Other Cognitive Disorders Unit, Neurology Service, Hospital Clínic, Institut d’Investigacións Biomèdiques August Pi I Sunyer, University of Barcelona, Barcelona, Spain; Cognitive Disorders Unit, Department of Neurology, Hospital Universitario Donostia, San Sebastian, Gipuzkoa, Spain; Neuroscience Area, Biodonostia Health Research Insitute, San Sebastian, Gipuzkoa, Spain; Clinique Interdisciplinaire de Mémoire, Département des Sciences Neurologiques, CHU de Québec, and Faculté de Médecine, Université Laval, Québec, Canada; Karolinska Institutet, Department NVS, Center for Alzheimer Research, Division of Neurogenetics, Stockholm, Sweden; Department of Neurodegenerative Diseases, Hertie-Institute for Clinical Brain Research & Center of Neurology, University of Tübingen, Germany; German Center for Neurodegenerative Diseases (DZNE), Tübingen, Germany; University of Milan, Centro Dino Ferrari, Milan, Italy; Fondazione IRCSS Ca’ Granda, Ospedale Maggiore Policlinico, Neurodegenerative Diseases Unit, Milan, Italy; LC Campbell Cognitive Neurology Research Unit, Sunnybrook Research Institute, Toronto, Ontario, Canada; Toronto Western Hospital, Tanz Centre for Research in Neurodegenerative Disease, Toronto, Ontario, Canada; Department of Clinical Neurological Sciences, University of Western Ontario, London, ON, Canada; Laboratory for Cognitive Neurology, Department of Neurosciences, KU Leuven, Leuven, Belgium; Neurology Service, University Hospitals Leuven, Belgium, Laboratory for Neurobiology, VIB-KU; Laboratory of Neurosciences, Institute of Molecular Medicine, Faculty of Medicine, University of Lisbon, Lisbon, Portugal; Fondazione Istituto di Ricovero e Cura a Carattere Scientifico Istituto Neurologico Carlo Besta, Milan, Italy; Neurology Department, Centro Hospitalar e Universitário de Coimbra, Coimbra, Portugal; Faculty of Medicine, University of Coimbra, Coimbra, Portugal; Centre of Neurosciences and Cell biology, Universidade de Coimbra, Coimbra, Portugal; Department of Psychiatry, McGill University Health Centre, McGill University, Montreal, Canada; McConnell Brain Imaging Centre, Montreal Neurological Institute, McGill University, Montreal, Canada; Nuffield Department of Clinical Neurosciences, Medical Sciences Division, University of Oxford, Oxford, UK; Division of Neuroscience and Experimental Psychology, Wolfson Molecular Imaging Centre, University of Manchester, Manchester, UK; Departments of Geriatric Medicine and Nuclear Medicine, University of Duisburg-Essen, Germany; Neurologische Klinik und Poliklinik, Ludwig-Maximilians-Universität, Munich, German Center for Neurodegenerative Diseases (DZNE), Munich, Germany; Department of Neurology, University Hospital Ulm, Ulm, Germany; Istituto di Ricovero e Cura a Carattere Scientifico (IRCCS) Istituto Centro San Giovanni di Dio Fatebenefratelli, Brescia, Italy; Memory Clinic and LANVIE-Laboratory of Neuroimaging of Aging, University Hospitals and University of Geneva, Geneva, Switzerland; Molecular Markers Laboratory, IRCCS Istituto Centro San Giovanni di Dio Fatebenefratelli, Brescia, Italy; Department of Neuroscience, Psychology, Drug Research and Child Health, University of Florence, Florence, Italy; Istituto di Ricovero e Cura a Carattere Scientifico (IRCCS) “Don Gnocchi”, Florence, Italy; Dementia Research Centre, Department of Neurodegenerative Disease, UCL Institute of Neurology, Queen Square, London, UK

**Keywords:** frontotemporal dementia (FTD), presymptomatic, functional magnetic resonance imaging (*f*MRI), network connectivity

## Abstract

**INTRODUCTION:** The presymptomatic phase of neurodegenerative disease can last many years, with sustained cognitive function despite progressive atrophy. We investigate this phenomenon in familial Frontotemporal dementia (FTD).

**METHODS:** We studied 121 presymptomatic FTD mutation carriers and 134 family members without mutations, using multivariate data-driven approach to link cognitive performance with both structural and functional magnetic resonance imaging. Atrophy and brain network connectivity were compared between groups, in relation to the time from expected symptom onset.

**RESULTS:** There were group differences in brain structure and function, in the absence of differences in cognitive performance. Specifically, we identified behaviourally-relevant structural and functional network differences. Structure-function relationships were similar in both groups, but coupling between functional connectivity and cognition was stronger for carriers than for non-carriers, and increased with proximity to the expected onset of disease.

**DISCUSSION:** Our findings suggest that maintenance of functional network connectivity enables carriers to maintain cognitive performance.

## 1. Introduction

Across the adult healthy lifespan, the structural and functional properties of brain networks are coupled, and both are predictive of cognitive ability [1,2]. The connections between structure, function and performance have been influential in developing current models of ageing and neurodegeneration [3–5]. However, this work contrasts with the emerging evidence of neuropathological and structural changes many years before the onset of symptoms of Alzheimer’s disease and frontotemporal dementia (FTD) [6–8]. Genetic FTD with highly-penetrant gene mutations provides the opportunity to examine the precursors of symptomatic disease. Three main genes account for 10-20% of FTD cases: chromosome 9 open reading frame 72 (*C9orf72*), granulin (*GRN*) and microtubule-associated protein tau (*MAPT*). These genes vary in their phenotypic expression and in the age of onset [9]. Despite pleiotropy [10], environmental and secondary genetic moderation [11,12] all three mutations cause significant structural brain changes in key regions over a decade before the expected age of disease onset [7,13], confirmed by longitudinal studies [14,15].

The divergence between early structural change and late cognitive decline begs the question: how do presymptomatic mutation carriers stay so well in the face of progressive atrophy? We propose that the answer lies in the maintenance of network dynamics and functional organisation [16]. Across the lifespan, functional brain network connectivity predicts cognitive status [17], and this connectivity-cognition relationship becomes stronger with age [18–20].

Our overarching hypothesis is that for those at genetic risk of dementia, the maintenance of network connectivity prevents the manifestation of symptoms despite progressive structural changes. A challenge is that neither the anatomical and functional substrates of cognition nor the targets of neurodegenerative disease are mediated by single brain regions: they are distributed across multi-level and interactive networks. We therefore used a multivariate data-driven approach to identify differences in the multidimensional brain-behaviour relationship between presymptomatic carriers and non-carriers of mutations in FTD genes. We identified key brain networks [21] from a large independent population-based age-matched dataset [22].

We tested three key hypotheses: (i) presymptomatic carriers differ from non-carriers in brain structure and brain function, but not in cognitive function, (ii) brain structure and function correlate with performance in both groups, but functional network indices are stronger predictors of cognition in carriers, and (iii) the dependence on network integrity for maintaining cognitive functioning increases as carriers approach the onset of symptoms.

## 2. Methods

### 2.1. Participants

Thirteen research sites across Europe and Canada recruited participants as part of an international multicentre partnership, the Genetic Frontotemporal Initiative (GENFI). 313 participants had usable structural and resting state functional magnetic resonance imaging data (MRI) [7,13]. The study was approved by the institutional review boards for each site, and participants providing written informed consent. Inclusion criteria included anyone over the age of 18, who is symptomatic or a an asymptomatic first-degree relative. Five participants were excluded due to excessive head motion (see below), resulting in 308 datasets for further analysis.

Participants were genotyped based on whether they carried a pathogenic mutation in *MAPT, GRN* and C9orf72. Mutation carriers were classified as either symptomatic or presymptomatic based on clinician evaluation. Participants were only classified as symptomatic if the clinician judged that symptoms were present, consistent with a diagnosis of a degenerative disorder, and progressive in nature. Additional group of controls, termed non-carriers, comprised of mutation-negative family members. In this study, we focus on non-carriers (NC, N=134) and presymptomatic carriers (PSC, N=121). Participants and site investigators were blinded to the research genotyping, although a minority of participants had undergone predictive testing outwith the GENFI study. See Table 1 for demographic information and Table 2 for behavioural, cognitive and neuropsychological information of both groups. In keeping with other GENFI reports, the years to expected onset (EYO) were calculated as the difference between age at assessment and mean age at onset within the family [7].

**Table 1.** Demographics of participants included in the analysis, grouped by genetic status as non-carriers (NC) and presymptomatic carriers (PSC). * denotes whether demographics vary between NC and PSC groups.

|  | Gene Status Group |  | Statistical tests* |  |
| --- | --- | --- | --- | --- |
|  | NC | PSC | X<sup>2</sup> or F-test | P-value |
| N | 134 | 121 |  |  |
| **Mutated gene, n (%)** |  |  | 0.86 | 0.649 |
| *MAPT* | 17 (12.7) | 19 (15.7) |  |  |
| *GRN* | 77 (57.5) | 63 (52.1) |  |  |
| *C9orf72* | 40 (29.9) | 39 (32.2) |  |  |
| **Gender, n (%)** |  |  | 0.01 | 0.908 |
| Male | 53 (39.6) | 47 (38.8) |  |  |
| **Handedness, n (%)** |  |  | 0.06 | 0.806 |
| Right-handed | 122 (91) | 107 (88.4) |  |  |
| **Age (Years)** |  |  | 2.68 | 0.103 |
| Mean / SD | 49 / 14 | 46 / 11 |  |  |
| Range [Min/Max] | 19 / 86 | 20 / 70 |  |  |
| **Expected Years to Onset** |  |  | 0.23 | 0.631 |
| Mean / SD | -10 / 12 | -11 / 11 |  |  |
| Range [Min/Max] | -25 / 10 | -25 / 10 |  |  |
| **Education (Years)** |  |  | 0.05 | 0.826 |
| Mean / SD | 14 / 3 | 14 / 3 |  |  |
| Range [Min/Max] | 5 / 24 | 5 / 22 |  |  |

\* Statistical test to indicate whether demographics vary between NC and PSC groups

**Table 2.**
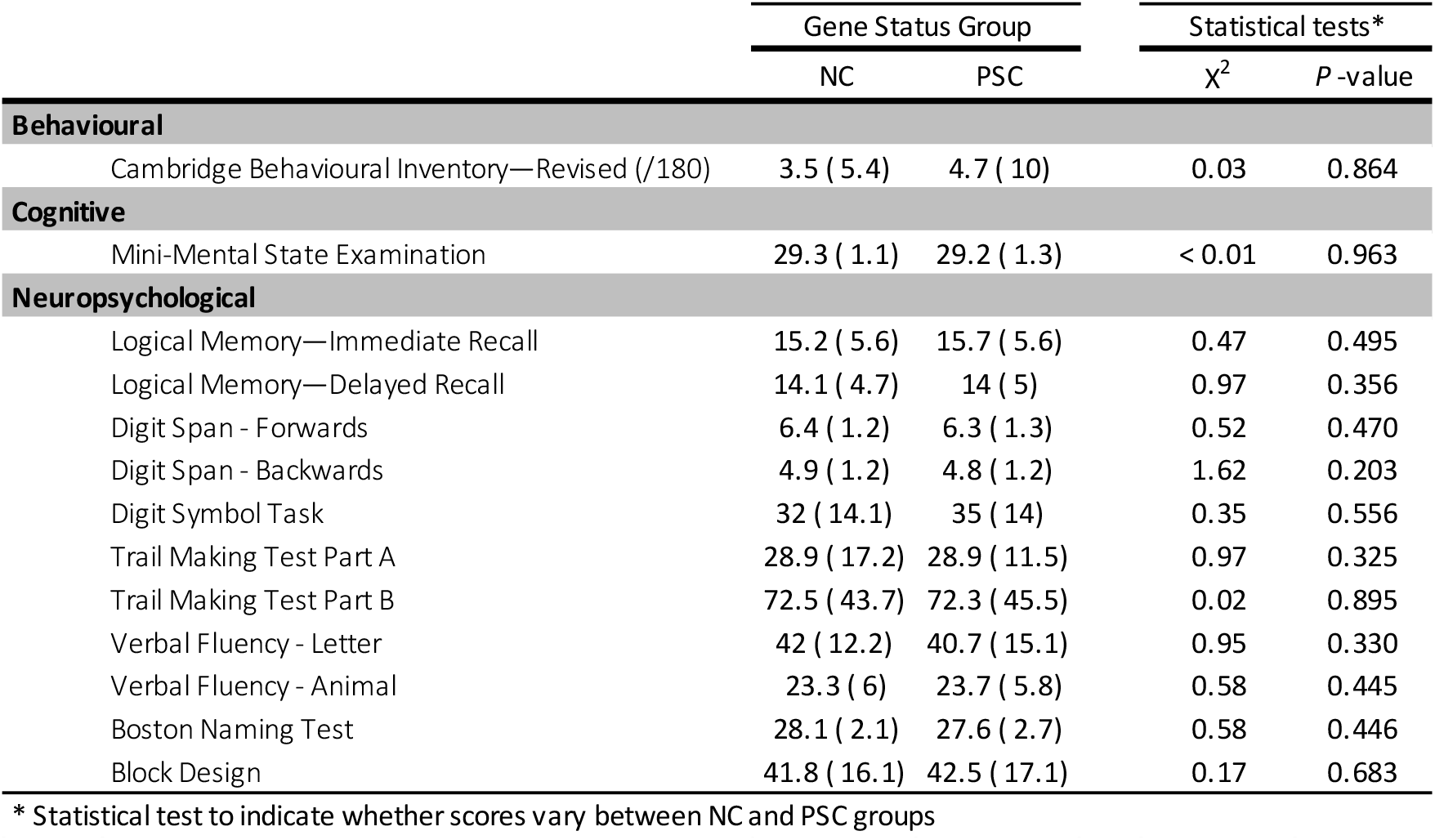
Behavioural, cognitive and neuropsychological estimates in presymptomatic carriers and non-carriers

|  | Gene Status Group |  | Statistical tests* |  |
| --- | --- | --- | --- | --- |
| | NC | PSC | $\chi^2$ | P-value |
| **Behavioural** |  |  |  |  |
| Cambridge Behavioural Inventory—Revised (/180) | 3.5 ( 5.4) | 4.7 ( 10) | 0.03 | 0.864 |
| **Cognitive** |  |  |  |  |
| Mini-Mental State Examination | 29.3 ( 1.1) | 29.2 ( 1.3) | < 0.01 | 0.963 |
| **Neuropsychological** |  |  |  |  |
| Logical Memory—Immediate Recall | 15.2 ( 5.6) | 15.7 ( 5.6) | 0.47 | 0.495 |
| Logical Memory—Delayed Recall | 14.1 ( 4.7) | 14 ( 5) | 0.97 | 0.356 |
| Digit Span - Forwards | 6.4 ( 1.2) | 6.3 ( 1.3) | 0.52 | 0.470 |
| Digit Span - Backwards | 4.9 ( 1.2) | 4.8 ( 1.2) | 1.62 | 0.203 |
| Digit Symbol Task | 32 ( 14.1) | 35 ( 14) | 0.35 | 0.556 |
| Trail Making Test Part A | 28.9 ( 17.2) | 28.9 ( 11.5) | 0.97 | 0.325 |
| Trail Making Test Part B | 72.5 ( 43.7) | 72.3 ( 45.5) | 0.02 | 0.895 |
| Verbal Fluency - Letter | 42 ( 12.2) | 40.7 ( 15.1) | 0.95 | 0.330 |
| Verbal Fluency - Animal | 23.3 ( 6) | 23.7 ( 5.8) | 0.58 | 0.445 |
| Boston Naming Test | 28.1 ( 2.1) | 27.6 ( 2.7) | 0.58 | 0.446 |
| Block Design | 41.8 ( 16.1) | 42.5 ( 17.1) | 0.17 | 0.683 |

\* Statistical test to indicate whether scores vary between NC and PSC groups

### 2.2. Neurocognitive assessment

Each participant completed a standard clinical assessment consisting of medical history, family history, functional status and physical examination, in complement with collateral history from a family member or a close friend. In the current study 13 behavioural measures of cognitive function were correlated with neuroimaging measures. These included the Uniform Data Set [23]: the Logical Memory subtest of the Wechsler Memory Scale-Revised with Immediate and Delayed Recall scores, Digit Span forwards and backwards from the Wechsler Memory Scale-Revised, a Digit Symbol Task, Parts A and B of the Trail Making Test, the short version of the Boston Naming Test, and Category Fluency (animals). Additional tests included Letter Fluency, Wechsler Abbreviated Scale of Intelligence Block Design task, and the Mini-Mental State Examination. Latency measures for the Trail Making Test were inverted so that higher values across all tests reflect better performance.

### 2.3. Neuroimaging assessment

Figure 1 provides a schematic representation of imaging data processing pipeline and the analysis strategy for linking brain-behaviour data. MRI data were acquired using 3T scanners and 1.5T where no 3T scanning was available from various vendors, with optimised scanning protocols to maximise synchronisation across scanners and sites [7,13]. A 3D-structural MRI was acquired on each participant using T1-weighted Magnetic Prepared Rapid Gradient Echo (MPRAGE) sequence over at least 283s (283-462s) and had a median isotropic resolution of 1.1mm (1-1.3mm), repetition time of 2000ms (6.6-2400), echo time of 2.9ms (2.6-3.5ms), inversion time of 8ms (8-9ms), and field of view 256×256×208mm (192-256×192-256×192-208mm). The co-registered T1 images were segmented to extract probabilistic maps of 6 tissue classes: grey matter (GM), white matter (WM), cerebrospinal fluid (CSF), bone, soft tissue, and residual noise. The native-space GM and WM images were submitted to diffeomorphic registration to create equally represented gene-group template images [DARTEL; 24]. The templates for all tissue types were normalised to the Montreal Neurological Institute template using a 12-parameter affine transformation. The normalised images were smoothed using an 8-mm Gaussian kernel.

**Figure 1.**
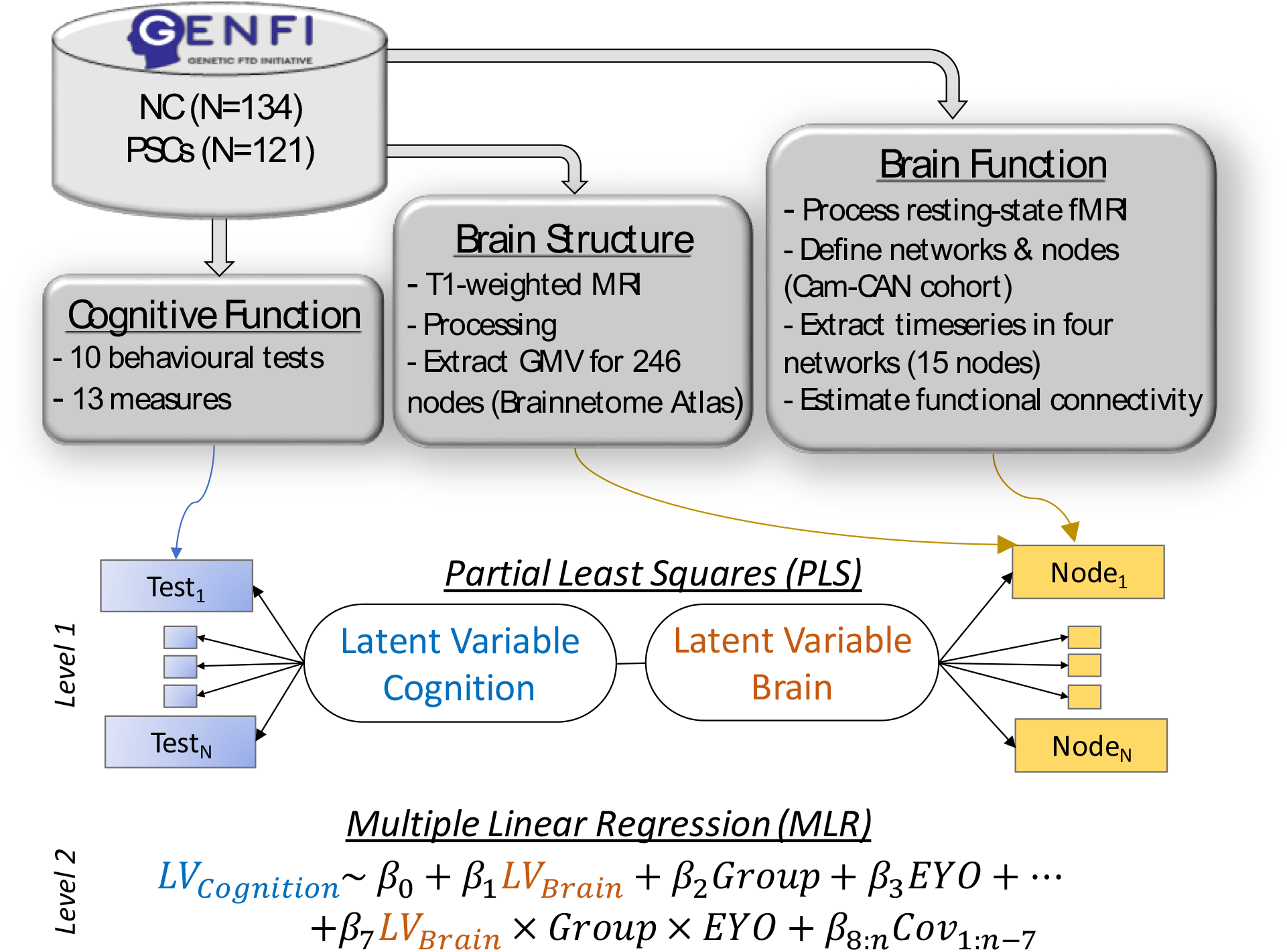
Schematic representation of data processing and analysis pipeline to test for brain-behaviour differences between presymptomatic carriers (PSC) and non-carriers (NC) as a function of expected years to onset (EYO) of symptoms, while controlling for covariates of no interest (Covs). Brain structural measures were based on the mean grey matter volume (GMV) in 246 nodes, as defined in the Brainnetome atlas [35]. Brain functional measures were based on the functional connectivity between 15 nodes as part of four large-scale networks, which were defined in an independent cohort of 298 age-matched individuals part of the Cam-CAN dataset.

For resting state fMRI measurements, Echo-Planar Imaging (EPI) data were acquired with at least six minutes of scanning. Analogous imaging sequences were developed by the GENFI Imaging Core team, and used at each GENFI study site to accommodate different scanner models and field strengths. EPI data were acquired over at least 300s (inter-quartile range 309-440) and had a median repetition time of 2200ms (2200-3000ms), echo time of 30ms, in-plane resolution of 2.75×2.75mm (2.75-3.31 x 2.75-3.31), and slice thickness of 3.3mm (3.0-3.3).

The imaging data were analysed using Automatic Analysis [AA 4.0, 25] pipelines and modules which called relevant functions from SPM12 [26]. To quantify the total motion for each participant, the root mean square volume-to-volume displacement was computed using the approach of Jenkinson et al [27]. Participants with 3.5 or more standard deviations above the group mean motion displacement were excluded from further analysis (N = 5). To further ensure that potential group bias in head motion did not affect later analysis of connectivity, we took three further steps: i) fMRI data was further postprocessed using whole-brain Independent Component Analysis (ICA) of single subject time-series denoising, with noise components selected and removed automatically using *a priori* heuristics using the ICA-based algorithm [28], ii) postprocessing of network node time-series (see below) and iii) a subject-specific estimate of head movement for each participant [27] included as a covariate in group-level analysis [29].

### 2.4. Network definition

The location of the key cortical regions in each network was identified by spatial-ICA in an independent dataset of 298 age-matched healthy individuals from a large population-based cohort [22]. Full details about preprocessing and node definition are described previously [30]. Four networks commonly affected by neurodegenerative diseases including FTD [21] were identified by spatially matching to pre-existing templates [31]. The node time-series were defined as the first principal component resulting from the singular value decomposition of voxels in an 8-mm radius sphere, which was centred on the peak voxel for each node [18]. Visual representation of the spatial distribution of the nodes is shown in Figure 2.

**Figure 2.**
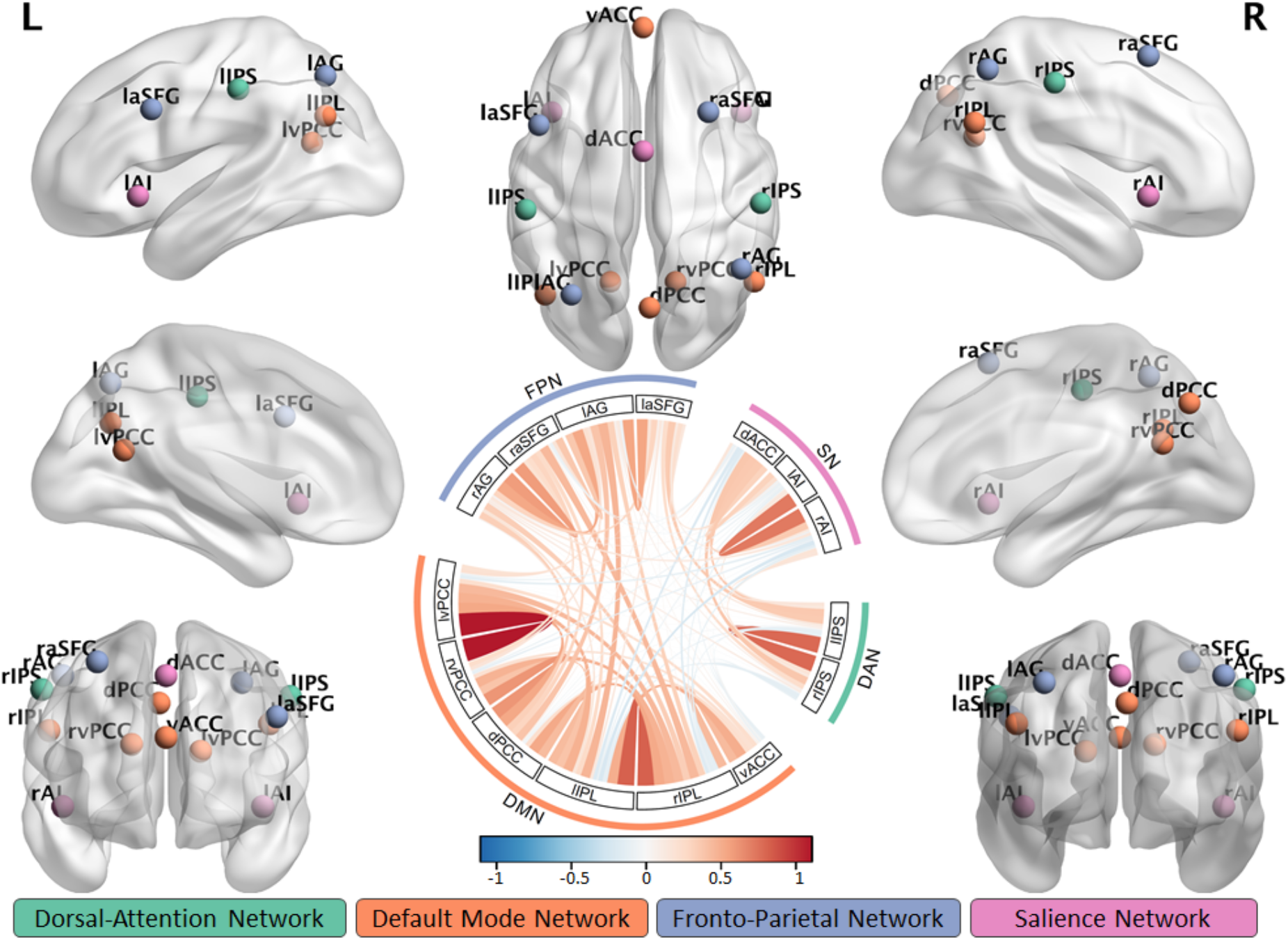
Visualisation of spatial localisation of the nodes part of the four large-scale networks and their mean functional connectivity (circular plot) across all participants in this study. Nodes and networks were defined in an independent cohort of 298 age-matched individuals part of the Cam-CAN dataset [30]. The default mode network (DMN) contained five nodes: the ventral anterior cingulate cortex (vACC), dorsal and ventral posterior cingulate cortex (vPCC and dPCC), and right and left inferior parietal lobes (rIPL and IIPL). The salience network (SN) was defined using right and left anterior insular (rAI and IAI) and dorsal anterior cingulate cortex (dACC). The frontoparietal network (FPN) was defined using right and left anterior superior frontal gyrus (raSFG and laSFG), and right and left angular gyrus (rAG and IAG). The dorsal attention Network (DAN) was defined using right and left intraparietal sulcus (rIPS and IIPS).

We aimed to further reduce the effects of noise confounds on functional connectivity effects of node time-series using general linear model (GLM) [29]. This model included linear trends, expansions of realignment parameters, as well as average signal in WM and CSF, including their derivative and quadratic regressors from the time-courses of each node. The WM and CSF signals were created by using the average signal across all voxels with corresponding tissue probability larger than 0.7 in associated tissue probability maps available in SPM12. A band-pass filter (0.0078-0.1 Hz) was implemented by including a discrete cosine transform set in the GLM. Finally, the functional connectivity (FC) between each pair of nodes was computed using Pearson’s correlation on postprocessed time-series.

### 2.5. Statistical analysis

#### 2.5.1. Group differences in brain structure, function and cognition

To assess the group-differences in neuroimaging and behavioural dataset we used multiple linear regression with a well-conditioned shrinkage regularization [32,33] and 10-Fold Cross–Validation [34]. In the analysis of brain structure we used as independent variables the mean grey matter volume (GMV) of the 246 brain nodes in the Brainnetome atlas [35]. The Brainnetome atlas was developed to link functional and structural characteristics of the human brain [35] and provides a fine-grained whole brain parcellation with a superior representation of age-related differences in brain structure compared to other cortical parcellation schemes [36,37]. In the analysis of brain function, we used the functional connectivity between 15 nodes, which were part of the four large-scale functional networks described above. In the analysis of cognitive function, the independent variables comprised the performance measures on the 13 neuropsychological tests performed outside of the scanner. In all three analyses the dependent variable was the genetic status (PSC vs NC) including age as a covariate of no interest. GENFI’s large-sampled cohort was created using harmonized multi-site neuroimaging data. Although, scanning protocols were optimised to maximise comparability across scanners and sites [7,13], different scanning platforms can introduce systematic differences which might confound true effects of interest [38]. Therefore, in the analysis of neuroimaging data we included scanner site and head motion as additional covariates of no interest.

#### 2.5.2. Brain-behaviour relationships

For the brain-behaviour analysis, we adopted a two-level procedure. In the first-level analysis, we assessed the multidimensional brain-behaviour relationships using partial least squares [39]. This analysis described the linear relationships between the two multivariate datasets, namely neuroimaging (either GMV or FC) and behavioural performance, by providing pairs of latent variables (Brain-LVs and Cognition-LVs) as linear combinations of the original variables which are optimised to maximise their covariance. Namely, dataset 1 consisted of a brain feature set, which could be either grey matter volume (GMV dataset) or functional connectivity strength between pairs of regions for each individual (FC dataset). Dataset 2 included the performance measures on the 13 tests (i.e. Cognition dataset), as considered in the multiple linear regression analysis of group differences in cognition. Covariates of no interest included head motion, scanner site, gender and handedness. In addition, we also included average GMV across all 15 nodes as a covariate of no interest in the FC-behaviour analysis to ensure that the observed effects are over and above differences in the level of atrophy.

Next, we tested whether the identified behaviourally-relevant LVs of brain structure and function were differentially expressed by NC and PSC as a function of expected years to onset. To this end, we performed a second-level analysis using multiple linear regression with robust fitting algorithm as implemented in matlab’s function “fitlm.m”. Independent variables included subjects’ brain scores from first level PLS (either Structure-LV or Function-LV subject scores), group information, expected years to onset and their interaction terms (e.g. brain scores x group, brain scores x years to expected onset, etc.). The dependent variable was subjects’ cognitive scores from the first level analysis in the corresponding PLS (Cognition-LV).

Given that the interaction effects were derived from continuous variables, we tested and interpreted interactions based on simple slope analysis and slope difference tests [40-42]. Covariates of no interest included gender, handedness, head movement and education (Figure 1). In addition, we included average GMV across all 15 nodes as a covariate in the FC-behaviour analysis to ensure that the observed effects are over and above differences in the level of atrophy.

## 3. Results

### 3.1. Group differences in neuroimaging and cognitive data

#### Brain structure

The multiple linear regression model testing for overall group differences in grey matter volume between PSC and NC was significant (r=.14, p=.025), reflecting expected presymptomatic differences in brain-wide atrophy. The frontal, parietal and subcortical regions had most atrophy in PSC (Figure 3). As expected, the group difference in grey matter volume of these regions increased as EYO decreased, see Supplementary Materials.

**Figure 3.**
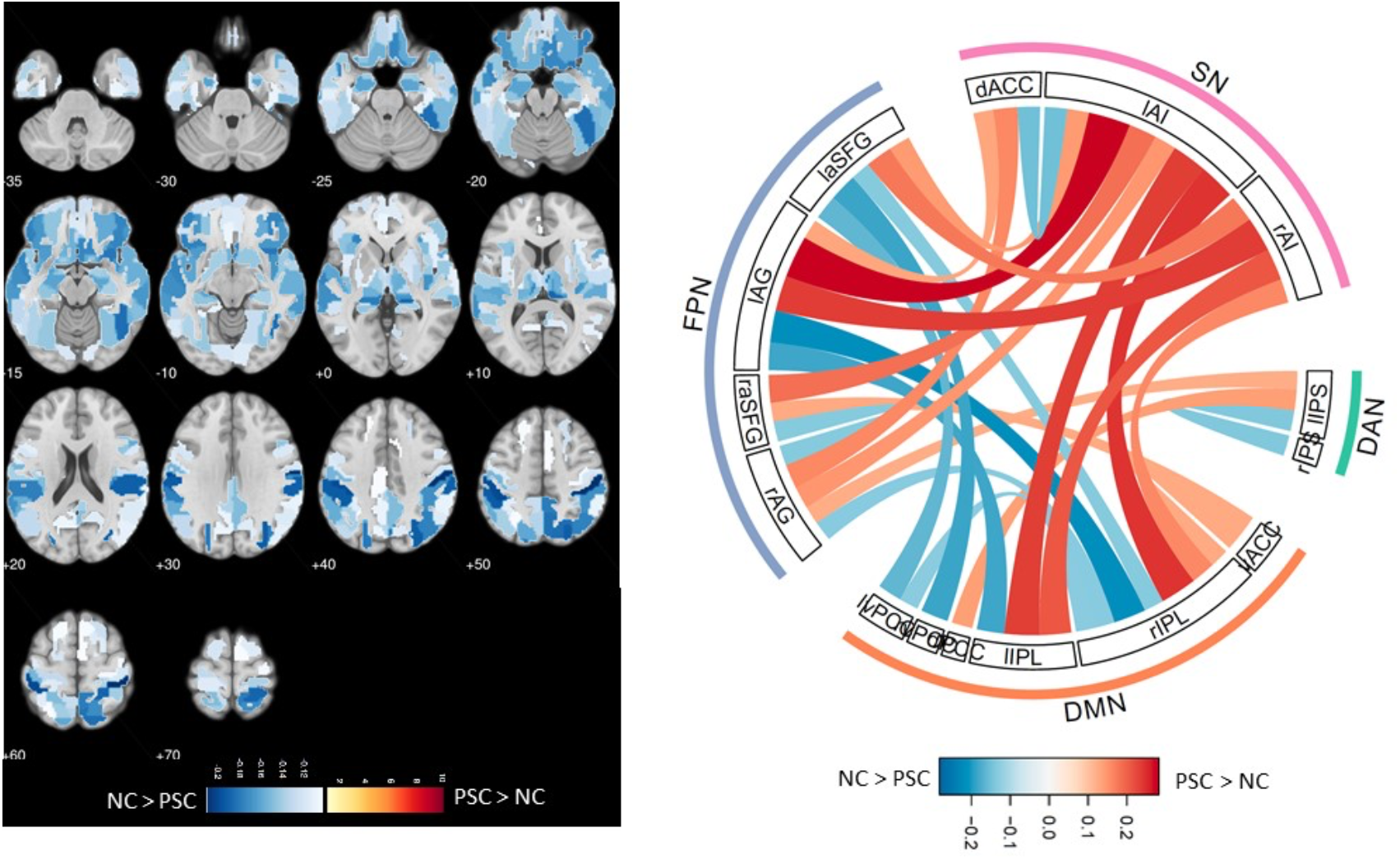
Group differences between PSC and NC in grey matter volume (left panel) and functional connectivity between nodes within four large scale networks (right panel). Hot colour scheme indicates the strength of effect size of PSC showing higher GMV and FC than NC, while cold colour scheme indicates the opposite effect (i.e. NC > PSC).

#### Brain Function

The multiple linear regression model testing for overall group differences in functional connectivity between PSC and NC was marginally significant (r=.12, p=.049). The pattern of connectivity indicated mainly increased connectivity between SN-DMN and SN-FPN in presymptomatic carriers, coupled with decreased connectivity within the networks and DMN-FPN connectivity (Figure 3).

#### Cognitive Function

We did not identify group differences in cognition and behaviour (r=.002, p=.807), confirming the impression of “healthy” status among presymptomatic carriers. However, in the next section, we consider the relationships between structure, function and cognition that underlie this maintenance of cognitive function.

### 3.2. Brain-behaviour relationships

#### Structure-cognition

Partial least squares analysis of grey matter volume and cognition identified one significant pair of latent variables (r = .40, p = .019). This volumetric latent variable expressed negative loadings in frontal (superior frontal gyrus, precentral gyrus, paracentral lobule), parietal (postcentral gyrus, precuneus, superior and inferior parietal lobule) and occipital (lateral and medial occipital cortex) regions and positive loadings in parahippocampal and hippocampal regions in addition to inferior temporal and insular cortex (Figure 4). The Cognition-LV profile expressed positively a large array of cognitive tests, with strongest values on delayed memory, Trail Making, Digit Symbol, Boston Naming and Fluency tests. The positive correlation between volumetric and cognitive LV’s confirms the expected relationship across the cohort as a whole, between cortical grey matter volume and both executive, language and mnemonic function (Figure 4).

**Figure 4.**
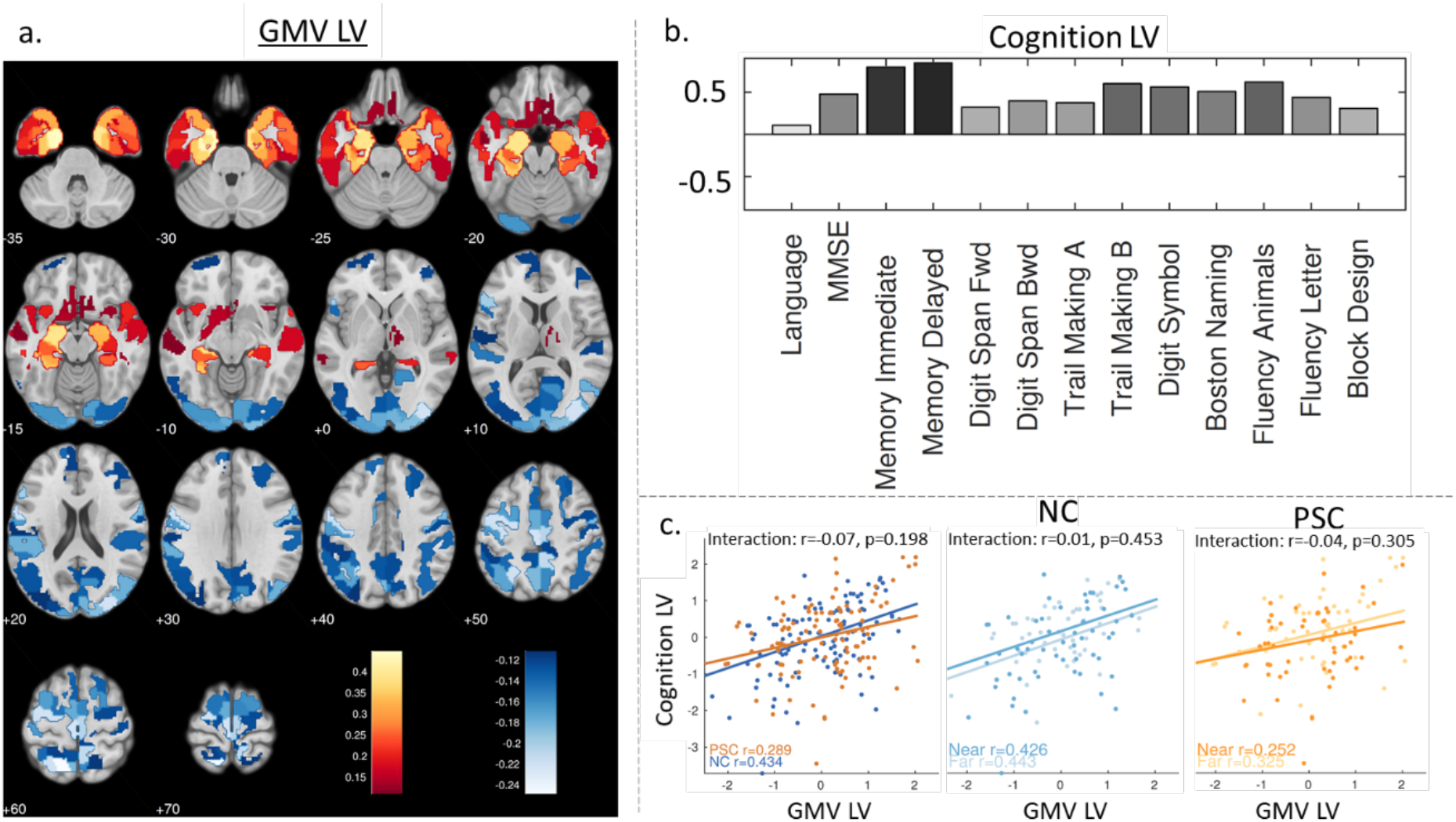
PLS analysis of grey matter volume (GMV) and cognition indicating the spatial distribution of GMV loading values (a), where hot and cold colour schemes are used for the strength of positive and negative correlations with the profile of Cognitive LV (b). (c) The scatter plot on the left represents the relationship between subjects scores of GMV LV and Cognition LV for presymptomatic carriers (PSC) and non-carriers (NC). The scatter plots in the middle and right hand-side represent GMV-Cognition LV relationship as a function of expected years to onset (EYO, split in two groups, Near and Far, see text) in each genetic status group separately.

To understand the structure-cognition relationship in each group and in relation to the expected years of onset, we performed a second-level interaction analysis using a regression model: we entered Cognition-LV subject scores as dependent variable, and grey matter volume LV subject scores, genetic status (i.e. mutation carrier or non-carrier), expected years to onset and their interactions as independent variables in addition to covariates of no interest. The results indicated that the relationship between grey matter volume and cognition could not be explained by genetic status, expected years to onset or their interactions with grey matter volume LV subject scores. There was no evidence for genetic status- and onset-dependent differences (over and above ageing and other covariates) in the associations between grey matter volume and cognition in this analysis (Figure 4).

#### Connectivity-Cognition

PLS analysis of functional connectivity and cognition also identified one significant pair of LVs (Function-LV and Cognition-LV, r=.32, p=.020), see Figure 5. This Function-LV reflected weak between-network connectivity, coupled with strong within-network connectivity. This pattern indicates the segregation or modularity of large-scale brain networks. The Cognition-LV expressed all tests, with positive loading values indicating that higher performance on a wide range of cognitive tests is associated with stronger functional network segregation. Cognitive deficits were associated with loss of segregation, with increased between-network connectivity and decreased within-network connectivity.

**Figure 5.**
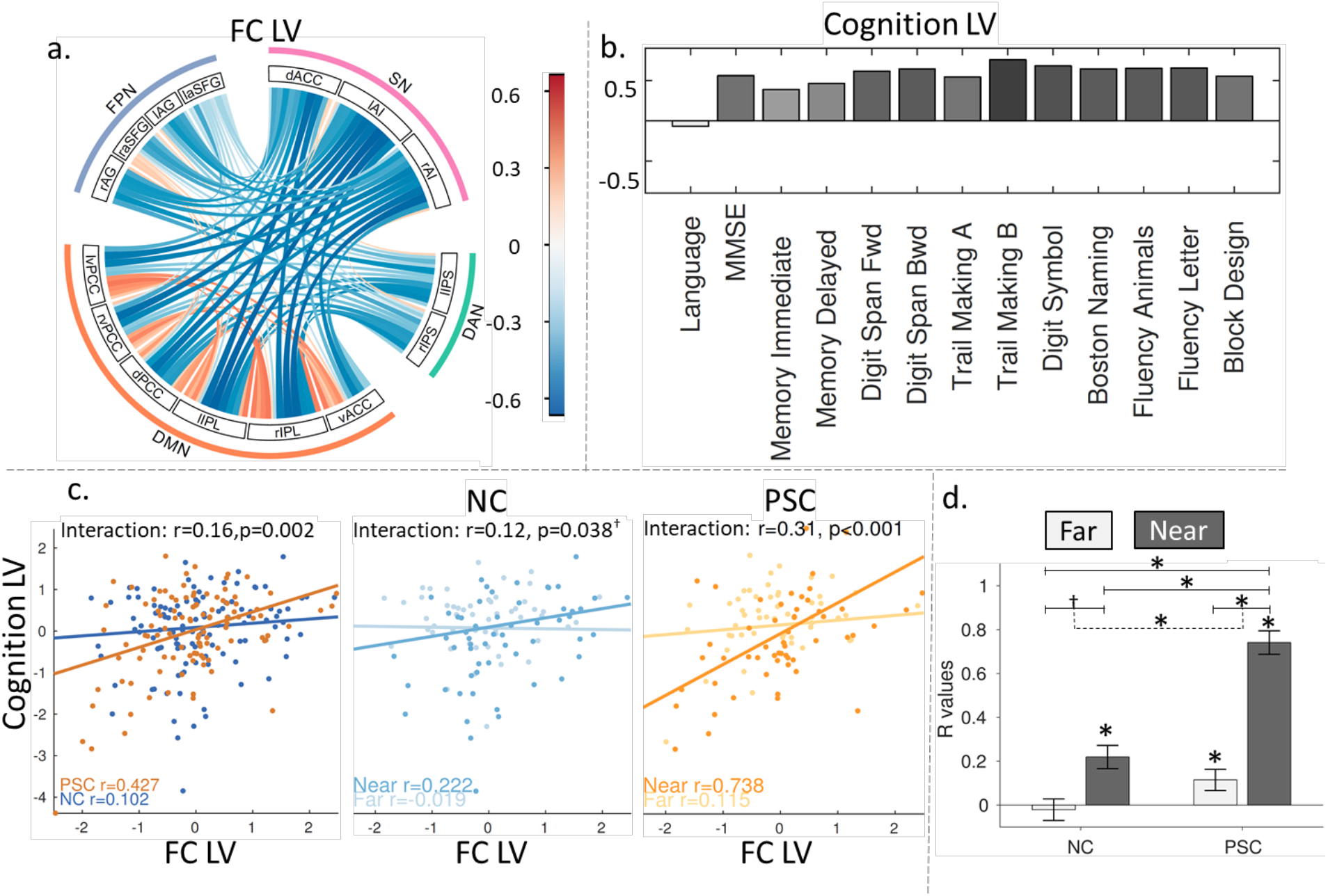
PLS analysis of functional connectivity and cognition indicating the connectivity pattern of loading values (a), where hot and cold colour schemes are used for the strength of positive and negative correlations with the profile of Cognitive LV (b). (c) The scatter plot on the left represents the relationship between subjects scores of Function LV and Cognition LV for presymptomatic carriers (PSC) and non-carriers (NC). The scatter plots in the middle and right hand-side represents Function-Cognition LV relationship as a function of expected years to onset (EYO split in two groups, Near and Far, see text) in each genetic status group separately. This is also represented using a bar chart in (d), where continuous and dashed lines indicate significance of effect differences and difference in differences, respectively. ^†^ and * denote significant tests at p-value < 0.05 (one- and two-sided, respectively).

To further test whether the observed behaviourally-relevant pattern of connectivity is differentially expressed between genetic status groups and expected years of onset, we constructed a second-level regression model with robust error estimates by including Function-LV subject scores, genetic status, expected years of onset and their interaction terms as independent variables and Cognition-LV as dependent variable in addition to covariates of no interest (Figure 5).

We found evidence for significant interaction between expected years of onset and Function-LV (r=.21, p<.001) and between group and Function-LV (r=.16, p=.002) explaining unique variance in Cognition-LV. We used simple slope analysis and slope difference tests [40–42] to test formally for differences in the relationship between Function-LV and Cognition-LV for PSC and NC. The relationship between Function-LV and Cognition-LV was stronger for PSC relative to NC (r=.16, p=.002), indicating the increasing importance of functional connectivity between the large-scale networks for PSC participants to maintain performance (Figure 5).

For ease of interpretation and illustration, we also computed the correlation between Cognition-LV and Function-LV for high and low levels of expected years to onset (EYO) within each group separately, where the levels were taken to be 1 standard deviation above and below the mean values of EYO following the simple slopes approach [40–42]. The two EYO subgroups were labelled “near” and “far”, with “near” for EYO values close to zero (i.e. participant’s age is “near” the age at which disease symptoms were demonstrated in the family), and “far” for EYO being a largely negative value (i.e. participant’s age is “far” from the age at which disease symptoms were demonstrated in the family). The analysis indicated that as the EYO decreases (i.e. participant’s age is reaching the years of onset of symptoms) the relationship between functional connectivity and performance becomes stronger. This effect was highly significant in presymptomatic carriers (r=.31, p<.001) and tended towards significance in non-carriers (r=.12, p=.038, one-sided). The differences in effects between presymptomatic carriers and non-carriers was qualified by a significant interaction term (t=2.27, p=0.024, i.e. the effect in presymptomatic mutation carriers was statistically stronger than the effect detected in non-carriers). These findings indicate that the relationship between FC and cognition is stronger in PSC relative to NC, and that this relationship increases as a function of EYO.

## 4. Discussion

In the present study, we confirmed previous findings of group differences in brain structure and function, in the absence of differences in cognitive performance between non-carriers and presymptomatic carriers of FTD-related genetic mutations. But, while the relationship between structure and cognition was similar in both groups, the coupling between function and cognition was stronger for presymptomatic carriers, and increased as they approached the expected onset of disease.

These results suggest that people can maintain good cognitive abilities and successful day-to-day functioning despite significant neuronal loss and atrophy. This disjunction between structure and function is a feature of healthy ageing, but we have shown that it also characterises presymptomatic FTD, over and above the age effects in their other family members, despite widespread progressive atrophy. The multivariate approach reveals two key findings: (i) presymptomatic carriers express stronger between-network and weaker within-network functional connectivity than age-matched non-carriers, and (ii) as carriers approach their estimated age of symptom onset, and atrophy becomes evident, the maintenance of good cognition is increasingly associated with sustaining balance of within- and between-network integration.

This balance of within- and between-network connectivity is characteristic of segregated and specialized network organization of brain systems. Such functional segregation varies with physiological ageing [17,18,43], with cognitive function [18] and in individuals at risk for Alzheimer’s disease [44]. Graph-theoretic quantification of network organisation confirms the relevance of modularity and efficiency to function in FTD [16]. Conversely, the loss of neural systems’ modularity mirrors the loss of functional specialization with age [45] and dementia [44]. Here, we show the significance of the maintenance of this functional network organisation, with a progressively stronger correlation with cognitive performance as seemingly healthy adults approach the age of expected onset of FTD.

The uncoupling of brain function from brain structure indicates that there may be independent and synergistic effects of multiple factors leading to cognitive preservation. This is consistent with a previous work in healthy ageing where brain activity and connectivity provide independent and synergistic predictions of performance across the lifespan [19]. Therefore, future studies need to consider the independent and synergistic effects of many possible biomarkers, based on MRI, computed tomography, positron-emission tomography, CSF, blood and brain histopathology. For example, functional network impairment may be related to tau expression and tau pathology, amyloid load, or neurotransmitter deficits in neurodegenerative diseases, independent of atrophy [30,46–48]. Importantly, studies need to recognise the rich multivariate nature of cognition and of neuroimaging in order to improve stratification procedures, e.g. based on integrative approaches that explain individual differences in cognitive impairment [30,49]. On a clinical level, this may facilitate future studies to establish whether presymptomatic carriers who maintain such connectivity profiles and thereby neuropsychological function in the presence of atrophy may have a lower risk of progression and better prognosis – information which will be important for future triallists, patients and carers.

We also recognise the difficulty to determine a unique contribution of each factor (e.g. brain structure and brain function), given the increasing interaction between factors in advanced stages of disease [50]. This is further complicated by these alterations becoming irreversible with progression of neurodegeneration [51]. This suggests that the critical interplay between multiple factors (including brain structure and function) may be better studied in the asymptomatic and preclinical stages as well as across the healthy lifespan, which could still be modifiable and their influences are likely to be more separable.

Our findings agree with the model of compensation in the presymptomatic and early phases of Huntington’s disease, where network coupling predicted better cognitive performance [52]. In a recent longitudinal study a non-linear concave-down pattern of both brain activity and behaviour was present, despite a linear decline in brain volume over time, [53]. Similar effects have been observed also in healthy ageing and amnestic mild cognitive impairment, where greater connectivity with the default-mode network and weaker connectivity between default-mode network and dorsal-attention network was associated with higher cognitive status in both groups [54]. Network integrity may also play a role in compensatory mechanisms in non-cognitive symptoms, such as motor impairment in Parkinson’s disease [55]. Accordingly, increased network efficiency and connectivity has been shown in prodromal phases, followed by decreased local connectivity in symptomatic phases, suggesting the emergence and dissipation of neural compensation [56].

The current study has several limitations. First, despite the large size of the overall GENFI cohort, we did not analyse each genetic group separately. The subdivision of each clinical group (PSC, NC) by three genes would have led to small and unbalanced subgroups, lowering statistical power and robustness. Moreover, genetic FTD is also characterised by multiple mutations within MAPT and GRN, and pleiotropy of clinical phenotypes from the same mutation [10]. Pleiotropy of clinical phenotype is avoided by the study of presymptomatic carriers, but we cannot rule out pleiotropy of intermediate phenotypes expressed as say neural network diversity. In FTD as in other dementias, clinical heterogeneity is modified by environmental factors such as education [which may be a surrogate of cognitive reserve, 12,57]. In addition, our analysis included the estimated age of onset in some models, but we recognise that the precision of the estimated years of onset (based on family history of onset) varies across mutations and families [7,58], being highest for MAPT and low for C9ORF72 expansion. Genetic modifiers such as TMEM106B [59], APOE [60], have also been identified. Further work, with larger cohorts is required to test for gene-specific effects, and the role of environmental and genetic moderators on the relationships between brain structure, functional networks and cognition. The harmonisation of sequences and data acquisition protocols in this multi-site neuroimaging study aimed to reduce the susceptibility to systematic differences across scanning platforms, but residual site variance cannot be ruled out [38,61]. The inclusion of study site as a covariate of no interest [61] and the nature of our multivariate approach to identify shared signals between brain and behavioural data reduce residual effects of scanner variance [38,62]. Future studies may use alternative brain measures that reflect differences in cortical surface and thickness estimates [63,64], or which infer neural connectivity directly from neurophysiology or from the separation of neurovascular from neuronal contributors to BOLD fMRI variance [18,65], given the confounding effects of age, drug or disease on neurovascular signals [66,67].

The current study is cross-sectional. Therefore, we cannot infer longitudinal progression within subjects as the unambiguous cause of the effects we observe in relation to expected years of onset. Accumulating evidence suggests that network integrity serves to maintain performance with either physiological ageing or pathological conditions. However, longitudinal mediation studies and pharmacological or electroceutical interventions would be needed to prove its causal role in cognitive preservation. Finally, our findings are limited to autosomal dominant FTD, which represents a minority of FTD: generalisation to sporadic forms of disease would be speculative.

In conclusion, we used a multivariate data-driven approach to demonstrate that brain functional integrity may facilitate presymptomatic carriers to maintain cognitive performance in the presence of progressive brain atrophy for years before the onset of symptoms. The multivariate approach to cognition and brain function is well-suited to address the effects of multiple interacting risk factors on biomarkers of the progression of neurodegeneration, ahead of clinical conversion to dementia. The approach and our findings have implications for the design of presymptomatic disease-modifying therapy trials, which are likely to rely initially on surrogate markers of brain health rather than clinical endpoints.

## Data Availability

Data access and sharing are subject to consent and GENFI consortium oversight. For enquires regarding data access please contact the senior author in the first instance.

## Acknowledgements

K.A.T. is supported by the British Academy Postdoctoral Fellowship (PF160048) and the Guarantors of Brain (101149). J.B.R. is supported by the Wellcome Trust (103838) the Medical Research Council (SUAG/051 G101400) and the Cambridge NIHR Biomedical Research Centre. R. S.-V. is supported by the Instituto de Salud Carlos III and the JPND network PreFrontAls (01ED1512/AC14/0013) and the Fundació Marató de TV3 (20143810). M.M and E.F are supported by the UK Medical Research Council, the Italian Ministry of Health and the Canadian Institutes of Health Research as part of a Centres of Excellence in Neurodegeneration grant, and also a Canadian Institutes of Health Research operating grant (MOP 327387) and funding from the Weston Brain Institute. J.D.R., D.C. and K.M.M. are supported by the NIHR Queen Square Dementia Biomedical Research Unit, the NIHR UCL/H Biomedical Research Centre and the Leonard Wolfson Experimental Neurology Centre (LWENC) Clinical Research Facility. J.D.R. is supported by an MRC Clinician Scientist Fellowship (MR/M008525/1) and has received funding from the NIHR Rare Disease Translational Research Collaboration (BRC149/NS/MH), the MRC UK GENFI grant (MR/ M023664/1) and The Bluefield Project. F.T. is supported by the Italian Ministry of Health (Grant NET-201102346784). L.C.J. and J.V.S. are supported by the Association for Frontotemporal Dementias Research Grant 2009, ZonMw Memorabel project number 733050103 and 733050813, and the Bluefield project. R.G. supported by Italian Ministry of Health, Ricerca Corrente. The Swedish contributors C.G., L.O. and C.A. were supported by grants from JPND Prefrontals Swedish Research Council (VR) 529-2014-7504, Swedish Research Council (VR) 2015-02926, Swedish Research Council (VR) 2018-02754, Swedish FTD Initiative-Schorling Foundation, Swedish Brain Foundation, Swedish Alzheimer Foundation, Stockholm County Council ALF, Karolinska Institutet Doctoral Funding and StratNeuro, Swedish Demensfonden, during the conduct of the study.

## 9. APPENDIX

### List of other GENFI consortium members

Sónia Afonso - Instituto Ciencias Nucleares Aplicadas a Saude, Universidade de Coimbra, Coimbra, Portugal Maria Rosario Almeida - Centre of Neurosciences and Cell Biology, Universidade de Coimbra, Coimbra, Portugal Sarah Anderl-Straub – Department of Neurology, Ulm University, Ulm, GermanyChristin Andersson - Department of Clinical Neuroscience, Karolinska Institutet, Stockholm, Sweden
Anna Antonell - Alzheimer’s disease and other cognitive disorders unit, Neurology Department, Hospital Clinic, Institut d’Investigacions Biomèdiques, Barcelona, Spain
Silvana Archetti - Biotechnology Laboratory, Department of Diagnostics, Spedali Civili Hospital, Brescia, Italy Andrea Arighi - Fondazione IRCSS Ca’ Granda, Ospedale Maggiore Policlinico, Neurodegenerative Diseases Unit, Milan, Italy
Mircea Balasa - Alzheimer’s disease and other cognitive disorders unit, Neurology Department, Hospital Clinic, Institut d’Investigacions Biomèdiques, Barcelona, Spain
Myriam Barandiaran - Neuroscience Area, Biodonostia Health Research Institute, Paseo Dr Begiristain sn, CP 20014, San Sebastian, Gipuzkoa, Spain
Nuria Bargalló - Radiology Department, Image Diagnosis Center, Hospital Clínic and Magnetic Resonance Image core facility, IDIBAPS, Barcelona, Spain
Robart Bartha - Department of Medical Biophysics, Robarts Research Institute, University of Western Ontario, London, Ontario, Canada
Benjamin Bender - Department of Diagnostic and Interventional Neuroradiology, University of Tuebingen, Tuebingen, Germany
Luisa Benussi - Istituto di Ricovero e Cura a Carattere Scientifico Istituto Centro San Giovanni di Dio Fatebenefratelli, Brescia, Italy
Valentina Bessi - Department of Neuroscience, Psychology, Drug Research, and Child Health, University of Florence, Florence, Italy
Giuliano Binetti - Istituto di Ricovero e Cura a Carattere Scientifico Istituto Centro San Giovanni di Dio Fatebenefratelli, Brescia, Italy
Sandra Black - LC Campbell Cognitive Neurology Research Unit, Sunnybrook Research Institute, Toronto, Canada Martina Bocchetta – Dementia Research Centre, Department of Neurodegenerative Disease, UCL Institute of Neurology, Queen Square London, UK
Sergi Borrego-Ecija - Alzheimer’s disease and other cognitive disorders unit, Neurology Department, Hospital Clinic, Institut d’Investigacions Biomèdiques, Barcelona, Spain
Jose Bras – Dementia Research Institute, Department of Neurodegenerative Disease, UCL Institute of Neurology, Queen Square, London, UK
Rose Bruffaerts - Laboratory for Cognitive Neurology, Department of Neurosciences, KU Leuven, Leuven, Belgium
Paola Caroppo - Fondazione Istituto di Ricovero e Cura a Carattere Scientifico Istituto Neurologico Carlo Besta, Milan, Italy
David Cash – Dementia Research Centre, Department of Neurodegenerative Disease, UCL Institute of Neurology, Queen Square, London, UK
Miguel Castelo-Branco - Neurology Department, Centro Hospitalar e Universitário de Coimbra, Instituto de Ciências Nucleares Aplicadas à Saúde (ICNAS), Coimbra, Portugal
Rhian Convery – Dementia Research Centre, Department of Neurodegenerative Disease, UCL Institute of Neurology, Queen Square, London, UK
Thomas Cope – Department of Clinical Neuroscience, University of Cambridge, Cambridge, UK Maura
Cosseddu – Centre for Neurodegenerative Disorders, Neurology Unit, Spedali Civili Hospital, Brescia, Italy
María de Arriba - Neuroscience Area, Biodonostia Health Research Institute, Paseo Dr Begiristain sn, CP 20014, San Sebastian, Gipuzkoa, Spain
Giuseppe Di Fede - Fondazione Istituto di Ricovero e Cura a Carattere Scientifico Istituto Neurologico Carlo Besta, Milan, Italy
Zigor Díaz - CITA Alzheimer, San Sebastian, Spain
Katrina M Moore – Dementia Research Centre, Department of Neurodegenerative Disease, UCL Institute of Neurology, Queen Square, London, UK
Diana Duro - Faculty of Medicine, Universidade de Coimbra, Coimbra, Portugal
Chiara Fenoglio - University of Milan, Centro Dino Ferrari, Milan, Italy
Camilla Ferrari - Department of Neuroscience, Psychology, Drug Research, and Child Health, University of Florence, Florence, Italy
Carlos Ferreira - Instituto Ciências Nucleares Aplicadas à Saúde, Universidade de Coimbra, Coimbra, Portugal
Catarina B. Ferreira - Faculty of Medicine, University of Lisbon, Lisbon, Portugal
Toby Flanagan – Faculty of Biology, Medicine and Health, Division of Neuroscience and Experimental Psychology, University of Manchester, Manchester, UK
Nick Fox – Dementia Research Centre, Department of Neurodegenerative Disease, UCL Institute of Neurology, Queen Square, London, UK
Morris Freedman - Division of Neurology, Baycrest Centre for Geriatric Care, University of Toronto, Toronto, Canada Giorgio Fumagalli - Fondazione IRCSS Ca’ Granda, Ospedale Maggiore Policlinico, Neurodegenerative Diseases Unit, Milan, Italy; Department of Neuroscience, Psychology, Drug Research and Child Health, University of Florence, Florence, Italy
Alazne Gabilondo - Neuroscience Area, Biodonostia Health Research Institute, Paseo Dr Begiristain sn, CP 20014, San Sebastian, Gipuzkoa, Spain
Roberto Gasparotti - Neuroradiology Unit, University of Brescia, Brescia, Italy
Serge Gauthier - Department of Neurology and Neurosurgery, McGill University, Montreal, Québec, Canada
Stefano Gazzina - Centre for Neurodegenerative Disorders, Neurology Unit, Department of Clinical and Experimental Sciences, University of Brescia, Brescia, Italy
Giorgio Giaccone - Fondazione Istituto di Ricovero e Cura a Carattere Scientifico Istituto Neurologico Carlo Besta, Milan, Italy
Ana Gorostidi - Neuroscience Area, Biodonostia Health Research Institute, Paseo Dr Begiristain sn, CP 20014, San Sebastian, Gipuzkoa, Spain
Caroline Greaves – Dementia Research Centre, Department of Neurodegenerative Disease, UCL Institute of Neurology, Queen Square London, UK
Rita Guerreiro – Dementia Research Institute, Department of Neurodegenerative Disease, UCL Institute of Neurology, London, UK
Carolin Heller – Dementia Research Centre, Department of Neurodegenerative Disease, UCL Institute of Neurology, Queen Square, London, UK
Tobias Hoegen - Department of Neurology, Ludwig-Maximilians-University of Munich, Munich, Germany
Begoña Indakoetxea - Cognitive Disorders Unit, Department of Neurology, Donostia University Hospital, Paseo Dr Begiristain sn, CP 20014, San Sebastian, Gipuzkoa, Spain
Vesna Jelic - Division of Clinical Geriatrics, Karolinska Institutet, Stockholm, Sweden
Lize Jiskoot - Department of Neurology, Erasmus Medical Center, Rotterdam, The Netherlands
Hans-Otto Karnath - Section of Neuropsychology, Department of Cognitive Neurology, Center for Neurology &
Hertie-Institute for Clinical Brain Research, Tübingen, Germany
Ron Keren - University Health Network Memory Clinic, Toronto Western Hospital, Toronto, Canada
Maria João Leitão - Centre of Neurosciences and Cell Biology, Universidade de Coimbra, Coimbra, Portugal
Albert Lladó - Alzheimer’s disease and other cognitive disorders unit, Neurology Department, Hospital Clinic, Institut d’Investigacions Biomèdiques, Barcelona, Spain
Gemma Lombardi - Department of Neuroscience, Psychology, Drug Research and Child Health, University of Florence, Florence, Italy
Sandra Loosli - Department of Neurology, Ludwig-Maximilians-University of Munich, Munich, Germany
Carolina Maruta - Lisbon Faculty of Medicine, Language Research Laboratory, Lisbon, Portugal
Simon Mead - MRC Prion Unit, Department of Neurodegenerative Disease, UCL Institute of Neurology, Queen Square, London, UK
Lieke Meeter - Department of Neurology, Erasmus Medical Center, Rotterdam, Netherlands
Gabriel Miltenberger - Faculty of Medicine, University of Lisbon, Lisbon, Portugal
Rick van Minkelen - Department of Clinical Genetics, Erasmus Medical Center, Rotterdam, The Netherlands
Sara Mitchell - LC Campbell Cognitive Neurology Research Unit, Sunnybrook Research Institute, Toronto, Canada
Benedetta Nacmias - Department of Neuroscience, Psychology, Drug Research and Child Health, University of Florence, Florence, Italy
Mollie Neason - Dementia Research Centre, Department of Neurodegenerative Disease, UCL Institute of Neurology, Queen Square, London, UK
Jennifer Nicholas – Department of Medical Statistics, London School of Hygiene and Tropical Medicine, London, UK
Linn Öijerstedt - Department of Geriatric Medicine, Karolinska Institutet, Stockholm, Sweden
Jaume Olives - Alzheimer’s disease and other cognitive disorders unit, Neurology Department, Hospital Clinic, Institut d’Investigacions Biomèdiques, Barcelona, Spain
Alessandro Padovani - Centre for Neurodegenerative Disorders, Neurology Unit, Department of Clinical and Experimental Sciences, University of Brescia, Brescia, Italy
Jessica Panman – Department of Neurology, Erasmus Medical Center, Rotterdam, The Netherlands
Janne Papma - Department of Neurology, Erasmus Medical Center, Rotterdam, The Netherlands
Irene Piaceri - Department of Neuroscience, Psychology, Drug Research and Child Health, University of Florence, Florence
Michela Pievani - Istituto di Ricovero e Cura a Carattere Scientifico Istituto Centro San Giovanni di Dio Fatebenefratelli, Brescia, Italy
Yolande Pijnenburg - VUMC, Amsterdam, The Netherlands
Cristina Polito - Department of Biomedical, Experimental and Clinical Sciences “Mario Serio”, Nuclear Medicine Unit, University of Florence, Florence, Italy
Enrico Premi - Stroke Unit, Neurology Unit, Spedali Civili Hospital, Brescia, Italy
Sara Prioni - Fondazione Istituto di Ricovero e Cura a Carattere Scientifico Istituto Neurologico Carlo Besta, Milan, Italy
Catharina Prix - Department of Neurology, Ludwig-Maximilians-University Munich, Germany
Rosa Rademakers - Department of Neurosciences, Mayo Clinic, Jacksonville, Florida, USA
Veronica Redaelli - Fondazione Istituto di Ricovero e Cura a Carattere Scientifico Istituto Neurologico Carlo Besta, Milan, Italy
Tim Rittman – Department of Clinical Neurosciences, University of Cambridge, Cambridge, UK
Ekaterina Rogaeva - Tanz Centre for Research in Neurodegenerative Diseases, University of Toronto, Toronto, Canada
Pedro Rosa-Neto - Translational Neuroimaging Laboratory, McGill University Montreal, Québec, Canada
Giacomina Rossi - Fondazione Istituto di Ricovero e Cura a Carattere Scientifico Istituto Neurologico Carlo Besta, Milan, Italy
Martin Rossor – Dementia Research Centre, Department of Neurodegenerative Disease, UCL Institute of Neurology, Queen Square, London, UK
Beatriz Santiago - Neurology Department, Centro Hospitalar e Universitário de Coimbra, Coimbra, Portugal
Elio Scarpini - University of Milan, Centro Dino Ferrari, Milan, Italy; Fondazione IRCSS Ca’ Granda, Ospedale Maggiore
Policlinico, Neurodegenerative Diseases Unit, Milan, Italy
Sonja Schönecker - Neurologische Klinik, Ludwig-Maximilians-Universität München, Munich, Germany
Elisa Semler – Department of Neurology, Ulm University, Ulm, Germany
Rachelle Shafei – Dementia Research Centre, Department of Neurodegenerative Disease, UCL Institute of Neurology, Queen Square, London, UK
Christen Shoesmith - Department of Clinical Neurological Sciences, University of Western Ontario, London, Ontario, Canada
Miguel Tábuas-Pereira - Centre of Neurosciences and Cell Biology, Universidade de Coimbra, Coimbra, Portugal Mikel Tainta - Neuroscience Area, Biodonostia Health Research Institute, Paseo Dr Begiristain sn, CP 20014, San Sebastian, Gipuzkoa, Spain
Ricardo Taipa - Neuropathology Unit and Department of Neurology, Centro Hospitalar do Porto - Hospital de Santo António, Oporto, Portugal
David Tang-Wai - University Health Network Memory Clinic, Toronto Western Hospital, Toronto, Canada
David L Thomas - Neuroradiological Academic Unit, UCL Institute of Neurology, London, UK
Hakan Thonberg - Center for Alzheimer Research, Division of Neurogeriatrics, Karolinska Institutet, Stockholm, Sweden
Carolyn Timberlake - University of Cambridge, Cambridge, UK
Pietro Tiraboschi - Fondazione Istituto di Ricovero e Cura a Carattere Scientifico Istituto Neurologico Carlo Besta, Milano, Italy
Philip Vandamme - Neurology Service, University Hospitals Leuven, Belgium; Laboratory for Neurobiology, VIB-KU Leuven Centre for Brain Research, Leuven, Belgium
Mathieu Vandenbulcke - Geriatric Psychiatry Service, University Hospitals Leuven, Belgium; Neuropsychiatry, Department of Neurosciences, KU Leuven, Leuven, Belgium
Michele Veldsman - University of Oxford, UK
Ana Verdelho - Department of Neurosciences, Santa Maria Hospital, University of Lisbon, Portugal
Jorge Villanua - OSATEK Unidad de Donostia, San Sebastian, Gipuzkoa, Spain
Jason Warren – Dementia Research Centre, Department of Neurodegenerative Disease, UCL Institute of Neurology, Queen Square, London, UK
Carlo Wilke - Hertie Institute for Clinical Brain Research, University of Tuebingen, Tuebingen, Germany
Ione Woollacott – Dementia Research Centre, Department of Neurodegenerative Disease, UCL Institute of
Neurology, Queen Square, London, UK
Elisabeth Wlasich - Neurologische Klinik, Ludwig-Maximilians-Universität München, Munich, Germany
Henrik Zetterberg - Department of Neurodegenerative Disease, UCL Institute of Neurology, London, UK
Miren Zulaica - Neuroscience Area, Biodonostia Health Research Institute, Paseo Dr Begiristain sn, CP 20014, San Sebastian, Gipuzkoa, Spain.

## Notes

### Competing Interest Statement

The authors have declared no competing interest.

